# Weight-Adjusted Waist Index and Its Association with Pulmonary Function in Children: 6-17 Years: Cross-Sectional Findings

**DOI:** 10.1101/2025.06.03.25328930

**Authors:** Wenjun Zhai, Yongfang Zhou, Jiangli Cheng, Xinyuan Yang, Guopeng Liang, Junya Zhou

**Affiliations:** Department of Respiratory Therapy, West China Hospital of Sichuan University, Chengdu, Sichuan Province, P. R. China; College of Nursing and Health, Henan University, Kaifeng, Henan Province, P. R. China

**Keywords:** Lung Function, Weight-adjusted Waist Index, National Health and Nutrition Examination Survey, Child Health, Obesity, Abdominal Fat, Central Obesity

## Abstract

**Background:** Excess adiposity has been linked to compromised lung function and elevated susceptibility to respiratory diseases which may lead to long-term lung deterioration. Conventional indicators of obesity, such as BMI and waist circumference, have limitations due to the so-called “obesity paradox.” The Weight-Adjusted Waist Index (WWI) is another way to assess central fat distribution. However, its relationship with lung function in children has not been explored to date.

**Objective:** To explore the correlation between WWI and lung function parameters among children aged 6 to 17.

**Methods:** Data from NHANES 2007-2012 were used for cross-sectional analysis. An adjusted multivariate regression framework weighted by the inclusion sample was used to assess the relationship between WWI and pulmonary function indices (FVC, FEV_1_, FEF25% -75%, PEF, and FEV_1_/FVC). Possible nonlinear relationship was assessed using curve fitting and split linear regression. Subgroup analyses were performed to assess differences between groups.

**Results:** This study involved 3,717 US children. Higher WWI was associated with lower FVC, FEV1, FEV1 25%-75% and PEF (β = -0.28, -0.26, -0.30, -0.30, -0.30, and -0.58).

The association between FEV1/FEVC and WWI was nearly significant (β = -0.06, P = 0.054). The breakpoints for nonlinear associations were 10.83, 10.70, 10.70, 11, and 10.79 (P < 0.001). Subgroup analyses showed stronger correlations for boys and L-shaped correlations for boys.

**Conclusion:** A nonlinear negative correlation between WWI and respiratory capacity metrics was found in US children, emphasizing the potential relevance of maintaining optimal WWI to respiratory health. Prospective cohort studies are warranted to validate these preliminary observations.

## Introduction

Based on the World Health Organization’s definition, overweight and obesity are considered as a group of disorders in which there is an inordinate accumulation of adipose tissue in the body which may have a negative impact on health((1).Specifically for individuals aged 5-19 years, the 2007 WHO Growth Reference establishes diagnostic thresholds using age-sex adjusted BMI z-scores: ≥+1 SD for overweight and ≥+2 SD for obesity. The global prevalence of obesity continues to increase, driven by a interaction involving behavioral patterns, genetic redisposition, and environmental determinants with children disproportionately affected (2,3).Epidemiologic evidence links obesity to numerous health risks, with children particularly vulnerable. Childhood obesity usually persists into adulthood, leading to long-term health burdens(4),and chronic diseases at an early age, such as adverse psychosocial consequences, circulatory system disease and type 2 diabetes(5).

A range of factors influence lung function in children, including biological factors, metabolic factors, environmental exposures, and lifestyle behaviors. Of these, obesity is an important and often underestimated determinant(6).Obesity impairs pulmonary function through multiple pathways(7), encompassing mechanical constraints-increased abdominal mass, limited diaphragmatic excursion, reduced chest wall compliance-and impaired ventilation It increases the risk of developing asthma (8)obstructive sleep apnea(9), and COPD exacerbations disease(10).Moreover, obesity promotes systemic(11–13)and chronic inflammation (14,15)as well as metabolic dysregulation(16), all of which further compromise respiratory function. Although commonly used, traditional metrics such as BMI may inadequately capture fat distribution, particularly central adiposity (17).

A new adiposity metric, WWI, which is derived from the quotient of waist circumference and the square root of total body mass, has recently emerged as a viable alternative to traditional obesity indicators(18). By incorporating central adiposity while minimizing the confounding effects of body mass, WWI offers a refined assessment of obesity. WWI has proven valuable in assessing central obesity and predicting obesity-related health risks, with studies showing strong correlations with conditions such as stroke(19), children bone density(20), developmental disorders,(21) and myopia(22).

Additionally, emerging research suggests that WWI may hold significant potential in assessing the influence of obesity on children’s pulmonary function. Prior research(23) has identified an inverse association between the pulmonary function and WWI in adults. However, this relationship remains under-demonstrated in the pediatric population. Our study explored the relationship between WWI and lung function in children with the aim of informing early prevention strategies for obesity-related respiratory disorders. The obtained evidence helps provide new insights into the body composition determinants of pulmonary dysfunction and contributes to determining whether WWI is a useful diagnostic tool for children’s preventative health care programs.

## Materials and Methods

### Source of data and study population

The National Health and Nutrition Examination Survey (NHANES), a population-based epidemiological program conducted by the National Center for Health Statistics, employs multistage probability sampling to monitor cardiometabolic risk factors and nutritional biomarkers in the U.S. non-institutionalized population. This biennial national survey integrates three standardized data collection modalities: Computer-assisted interviews capturing dietary patterns, systematic physical examinations measuring anthropometric parameters, and centralized laboratory analyses quantifying metabolic indicators. Utilizing a multistage, stratified, and clustered sampling strategy, NHANES ensures nationally representative estimates. This analytical framework incorporated sampling weight adjustments to address multistage probability sampling characteristics, enabling derivation of nationally representative estimates. The NCHS Ethics Review Board approved all data collection protocols with documented participant consent. As the investigation utilized anonymized public datasets from NHANES, institutional review board exemption was confirmed. Comprehensive methodological specifications, survey design, procedures and ethical approval are accessible through official documentation repositories: https://www.cdc.gov/nchs/nhanes.

NHANES enrolled 30,442 respondents across survey waves conducted between 2007 and 2012. Participants aged 6 to 17 years were selected for inclusion (N = 6,791). Exclusions were made for missing WWI data (N = 454), absent spirometry measurements (N = 607), and incomplete covariate information (N = 2,015). After applying the exclusion criteria, the final analysis included 3,717 children. Details are in **Fig 1**.

**Fig 1.**
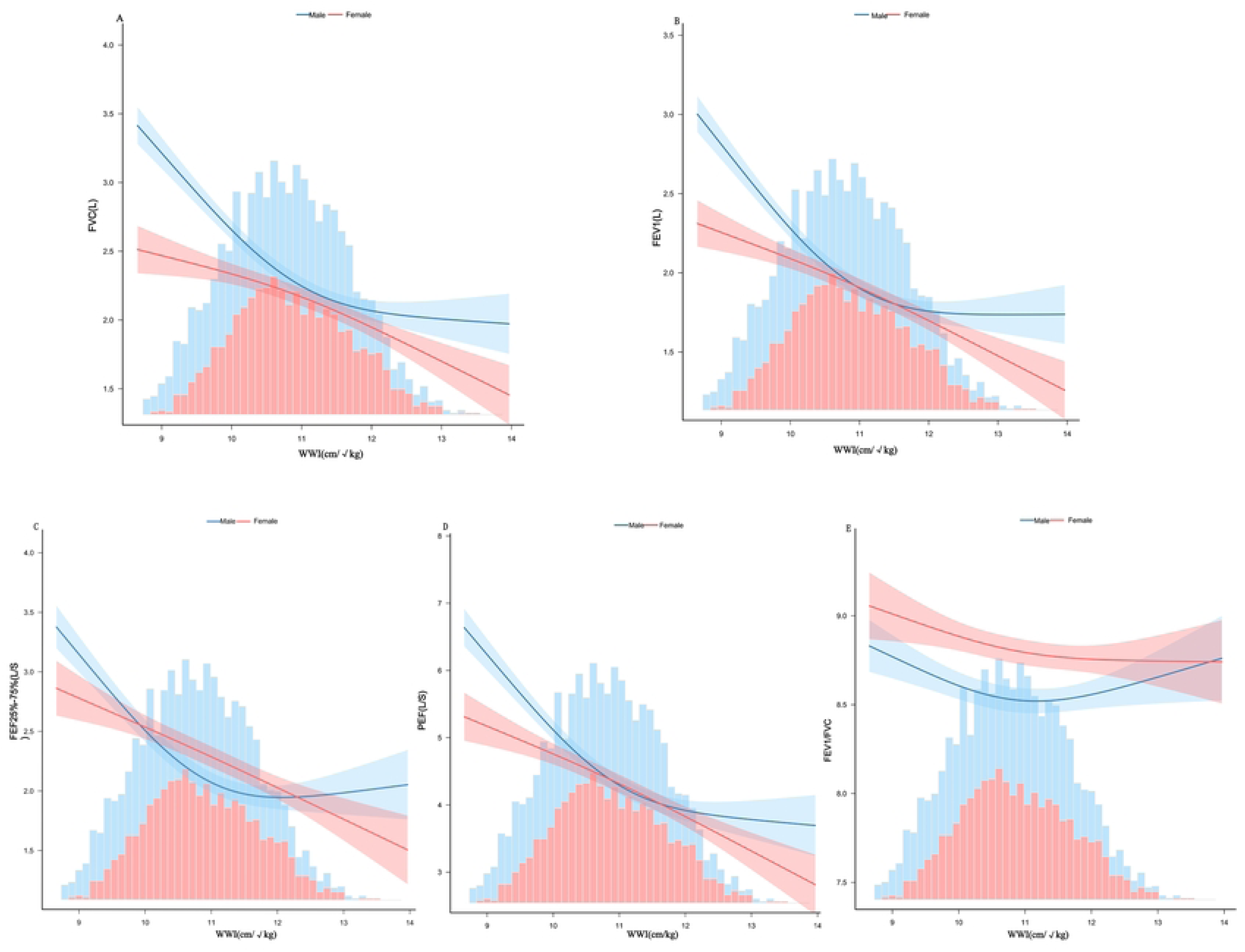
Flowchart of participant inclusion/exclusion criteria for weight-adjusted waist index (WWI) analysis using National Health and Nutrition Examination Survey (NHANES) data.

### WWI assessment

Certified examiners collected anthropometric measurements including waist circumference (WC, cm) and body weight (BW, kg) following NHANES protocols in mobile examination centers (MECs). Publicly accessible via the National Health and Nutrition Examination Survey repository, these data facilitated calculation of the weight-adjusted waist index (WWI) using the formula (18):

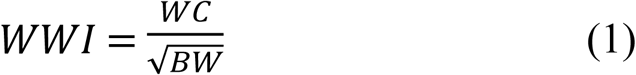

we operationalize WWI as the principal exposure variable and evaluated both as a continuous measure and as quartiles (Q1–Q4) to for subsequent analysis in this study.

### Lung function assessment

We extracted lung function data from the NHANES database for participants aged 6 to 17 years during the 2007–2012 cycles. We carried out pulmonary function tests in accordance with the American Thoracic Society (ATS).(24). Our exclusion standards were that the participant had current chest pains, physical limitations preventing maximal expiration, were receiving oxygen therapy, or had recently undergone surgery involving the chest, abdomen, or eyes. Additional exclusions included a recent heart attack, stroke, hemoptysis, or known tuberculosis exposure. Individuals with a history of retinal detachment or pneumothorax, and children with painful ear infections were also excluded. We looked at five key measures of lung function, namely forced vital capacity (FVC), forced expiratory volume in one second (FEV_1_), forced expiratory flow between 25% and 75% of lung volume (FEF25%–75%), peak expiratory flow (PEF), and the ratio of FEV_1_to FVC.

### Covariate assessment

Our analyses controlled a range of factors, including demographic factors such as racial or ethnic group, age, and gender; socioeconomic factors such as education level (25),citizenship category, and poverty-to-income ratio (PIR); and clinical factors including birth weight, serum cotinine levels(21),and family history of asthma(26).Educational attainment was stratified into three categories: less than grade 6, grades 6 through 9, and above grade 9.

PIR was grouped as follows: low (≤1.3), middle (1.3–3.5), and high (≥3.5). Serum cotinine levels served as a tobacco exposure biomarker and classified into three levels: no exposure (below the detection threshold of 0.05 ng/mL), secondhand exposure (between 0.05 and 2.00 ng/mL), and active exposure (2 ng/mL or higher2 ng/mL).

All data were collected by trained personnel through structured interviews, clinical examinations, and laboratory assessments. Further details exist on the official NHANES website: https://www.cdc.gov/nchs/nhanes/index.htm

### Statistical analysis

All analyses used two-tailed tests, considering a P value below 0.05 as statistically significant. processing data and analysis of stats were performed with Free Statistics version 2.0 (27)alongside R programming language implementations. To ensure our findings accurately reflect the non-institutionalized U.S. civilian population, we applied NHANES sampling weights in our multivariate linear regression analysis. This accounts for the survey’s complex multistage, stratified, and clustered sampling design. We applied weights using stratification and clustering variables to ensure unbiased estimates, accounting for variations in selection probability and response rates. In compliance with NHANES analytical recommendations sampling weights for the pooled 2007–2012 cycles were adjusted by dividing the original 2-year weights by six. The regression models accounted for the complex survey design and weights, implemented using the R ’survey’ package. We categorized participants into quartiles based on their WWI values and compared demographic characteristics across quartiles using weighted analyses. Weighted t-tests were applied to continuous factors, and weighted chi-square tests to categorical ones. We used weighted multivariable linear regression models to examine the relationship between WWI and pulmonary function and applied smooth curves to evaluate potential nonlinear relationships.

To identify inflection points in nonlinear patterns, we applied two-piecewise linear regression and log-likelihood ratio tests. In addition, we performed Subgroup analyses and tests for interaction to determine whether the association between WWI and pulmonary function differed across predefined Strata varied by sex, race/ethnicity, Poverty Income Ratio, Blood cotinine concentration, Weight at birth and citizenship status. To visualize these differences, we also plotted smooth curves stratified by sex. For the main analysis, we developed three regression models: Model 1: Unadjusted. Model 2 (Basic): Adjusted for education, age, citizenship, sex, and race/ethnicity. Model 3 (Full): Additionally adjusted for PIR, serum cotinine concentration, weight at birth, and blood relative asthma.

## Results

### Participants’ baseline attributes

Participant demographics and five pulmonary function indicators are detailed in **Table 1**; the cohort included 51.5% boys and 48.5% girls. The participants’ WWI values were used to stratify them into four quartiles, with the respective ranges for the first to fourth quartiles being 8.65 cm/√kg–10.15 cm/√kg, 10.15 cm/√kg–10.79 cm/√kg, 10.79 cm/√kg–11.42 cm/√kg, and 11.42 cm/√kg–13.96 cm/√kg. Pulmonary function measures declined progressively with higher WWI quartiles (P < 0.001). Top-quartile participants tended to be younger, boy, <6th Grade, lower weight at birth, Non-Hispanic White, Citizen by birth or naturalization, PIR<=1.3, Serum cotinine<0.05ng/ml, and No blood relatives have asthma. **(Table 1)**

**Table 1.** Description at baseline of the pediatric study population, grouped by WWI quartile.

| Variables | Weight-adjusted waist index (cm/$\sqrt{\text{kg}}$) | | | | | P-value |
| --- | --- | --- | --- | --- | --- | --- |
|  | Overall | Q1 | Q2 | Q3 | Q4 |  |
|  |  | (8.65–10.15) | (10.15–10.79) | (10.79–11.42) | (11.42–13.96) |  |
|  | N=28,137,362 | N=8,024,567 | N=7,181,195 | N=6,792,468 | N=6,139,130 |  |
| **WWI (cm/$\sqrt{\text{kg}}$)** |  |  |  |  |  |  |
| **(mean (SD))** | 10.71(0.866) | 9.68(0.347) | 10.48(0.180) | 11.08(0.182) | 11.91(0.394) | <0.001 |
| **SEX (%)** |  |  |  |  |  |  |
| **Male** | 51.49 | 60.34 | 43.88 | 50.33 | 50.12 | <0.001 |
| **female** | 48.51 | 39.66 | 56.12 | 49.67 | 49.88 |  |
| **AGE (years)** |  |  |  |  |  |  |
| **(mean (SD))** | 10.83(2.860) | 13.13(1.693) | 11.41(2.408) | 9.76(2.601) | 8.32(2.144) | <0.001 |
| **RACE (%)** |  |  |  |  |  |  |
| **Mexican American** | 14.79 | 7.69 | 14.18 | 16.44 | 22.96 | <0.001 |
| **Other Hispanic** | 6.81 | 5.64 | 6.93 | 6.70 | 8.31 |  |
| **Non-Hispanic White** | 57.83 | 58.50 | 58.82 | 59.41 | 54.02 |  |
| **Non-Hispanic Black** | 13.73 | 20.06 | 14.55 | 11.25 | 7.25 |  |
| **Other Race - Including Multi-Racial** | 6.84 | 8.11 | 5.52 | 6.20 | 7.46 |  |
| **Education (%)** |  |  |  |  |  |  |
| **>9th Grade** | 0.91 | 1.64 | 1.06 | 0.43 | 0.32 | <0.001 |
| **6th Grade-9th Grade** | 41.15 | 77.09 | 47.69 | 21.77 | 7.98 |  |
| **<6th Grade** | 57.94 | 21.27 | 51.25 | 77.80 | 91.70 |  |
| **PIR (%)** |  |  |  |  |  |  |
| **<=1.3** | 32.56 | 26.21 | 28.62 | 35.68 | 42.02 | <0.001 |
| **1.3-3.5** | 37.14 | 35.28 | 39.95 | 37.77 | 35.59 |  |
| **>=3.5** | 30.30 | 38.51 | 31.43 | 26.55 | 22.39 |  |
| **Birthweight, pounds** |  |  |  |  |  |  |
| **(mean (SD))** | 6.94(1.376) | 6.99(1.414) | 6.93(1.334) | 6.95(1.321) | 6.89(1.432) | 0.698 |
| **Serum cotinine(ng/ml)** |  |  |  |  |  |  |
| **<0.05** | 58.51 | 63.12 | 62.04 | 55.96 | 51.15 | 0.001 |
| **0.05-3** | 36.92 | 30.80 | 34.77 | 40.38 | 43.59 |  |
| **>=3** | 4.57 | 6.08 | 3.19 | 3.66 | 5.26 |  |
| **Blood relatives have asthma, n (%)** |  |  |  |  |  |  |
| **NO** | 69.05 | 71.11 | 68.97 | 68.85 | 66.66 | 0.466 |
| **YES** | 30.95 | 28.89 | 31.03 | 31.15 | 33.34 |  |
| **Citizenship (%)** |  |  |  |  |  |  |
| **Citizen by birth or naturalization** | 96.27 | 96.56 | 96.12 | 95.81 | 96.60 | 0.779 |
| Not a citizen of the US | 3.73 | 3.44 | 3.88 | 4.19 | 3.40 |  |
| FVC (L) (mean (SD)) | 2.84(1.048) | 3.63(0.905) | 2.97(0.942) | 2.47(0.891) | 2.07(0.688) | <0.001 |
| FEV<sub>1</sub>(L) (mean (SD)) | 2.44(0.899) | 3.13(0.774) | 2.54(0.811) | 2.10(0.747) | 1.77(0.562) | <0.001 |
| FEF<sub>25%-75%</sub>(L/S)<br>(mean (SD)) | 2.71(1.124) | 3.46(1.067) | 2.84(1.076) | 2.34(0.979) | 2.01(0.701) | <0.001 |
| PEF(L/S) (mean (SD)) | 5.51(1.936) | 7.02(1.612) | 5.71(1.717) | 4.83(1.644) | 4.04(1.281) | <0.001 |
| FEV<sub>1</sub>/FVC (mean (SD)) | 0.86(0.065) | 0.86(0.066) | 0.86(0.065) | 0.85(0.064) | 0.86(0.063) | 0.047 |

Continuous measurements are reported as mean (SD) and categorical as percentages. P values for continuous variables were derived from weighted linear regression analyses, whereas P values for categorical comparisons were obtained via weighted χ^2^ tests. Abbreviations: Q, quartile; PIR, poverty-to-income ratio; WWI, weight-adjusted waist index; FEV_1_, forced expiratory volume in 1 second; FVC, forced vital capacity; FEV_1_/FVC, ratio of FEV_1_to FVC; PEF, peak expiratory flow; FEF25%–75%, forced expiratory flow between 25% and 75% of FVC.

### Association between WWI and lung function indices

**Table 2** presents the association between WWI and pulmonary function parameters, based on weighted multiple linear regression analyses. In unadjusted models, higher WWI was inversely associated with FVC, FEF25%–75%, FEV_1_, and PEF (all P < 0.05), regardless of whether WWI was treated as a continuous or categorical variable. Upon adjustment for potential confounders, these negative associations persisted: FVC: β = –0.28; 95% CI, –0.32 to –0.24,FEV_1_: β = –0.26; 95% CI, –0.30 to –0.23, FEF25%–75%: β = –0.30; 95% CI, –0.36 to –0.25,PEF: β = –0.58; 95% CI, –0.66 to –0.51.The association between WWI and the ratio of FEV_1_/FVC approached statistical significance (β = –0.06; 95% CI, –0.06 to 0.00; P = 0.054).Analysis using WWI quartiles, with the lowest serving as the benchmark, showed a significant link between higher WWI levels and progressively lower FEV_1_, PEF, FVC, FEF25%–75% and FEV_1_/FVC values (P for trend < 0.05). These negative associations were also confirmed by non-linear analysis using smooth curve fitting **(Fig 2)**.

**Fig 2.**
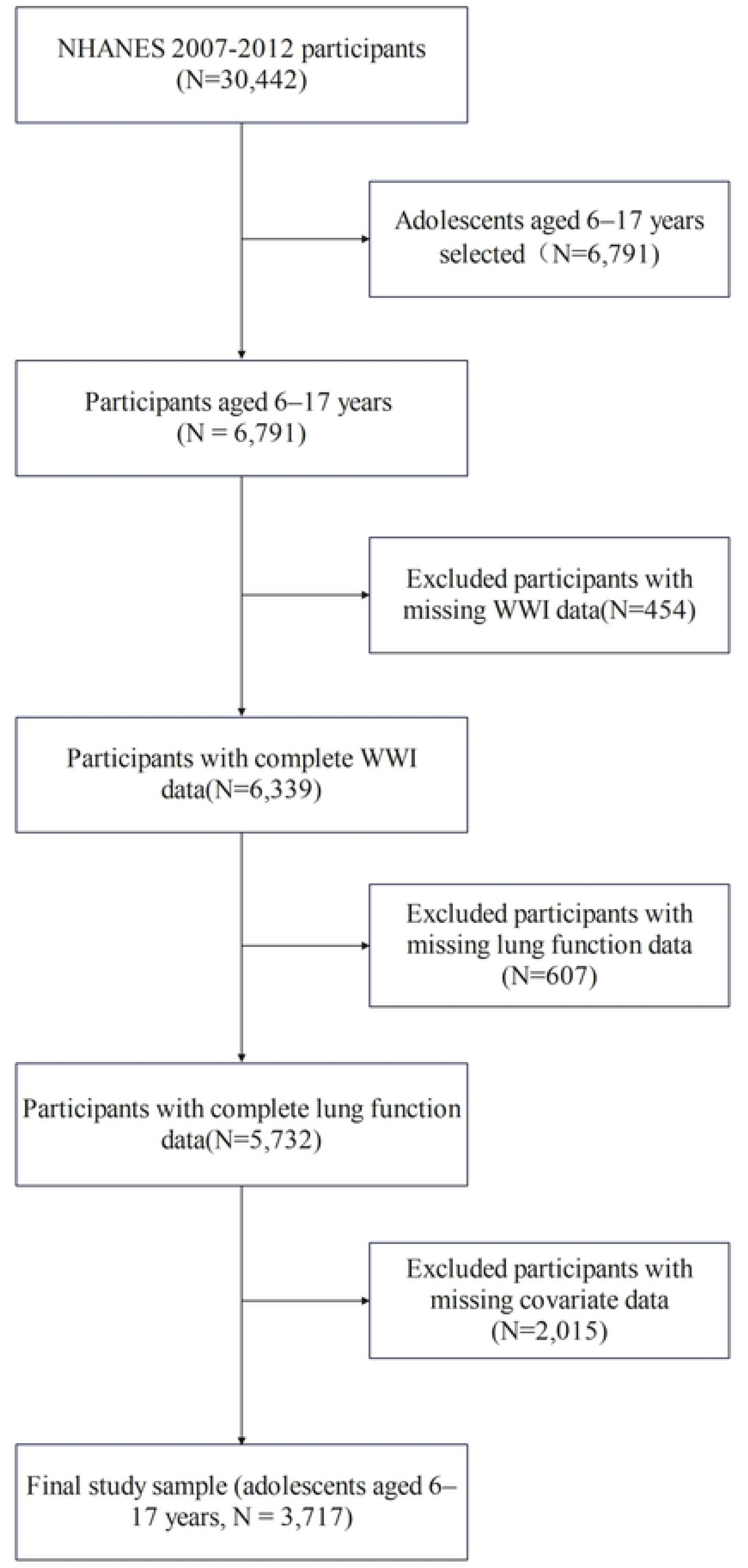
Exposure-Response Relationship between WWI and Pulmonary Function. Red solid lines represent the estimated association from a fully adjusted regression model, pink shaded areas indicate 95% confidence intervals.

**Table 2.**
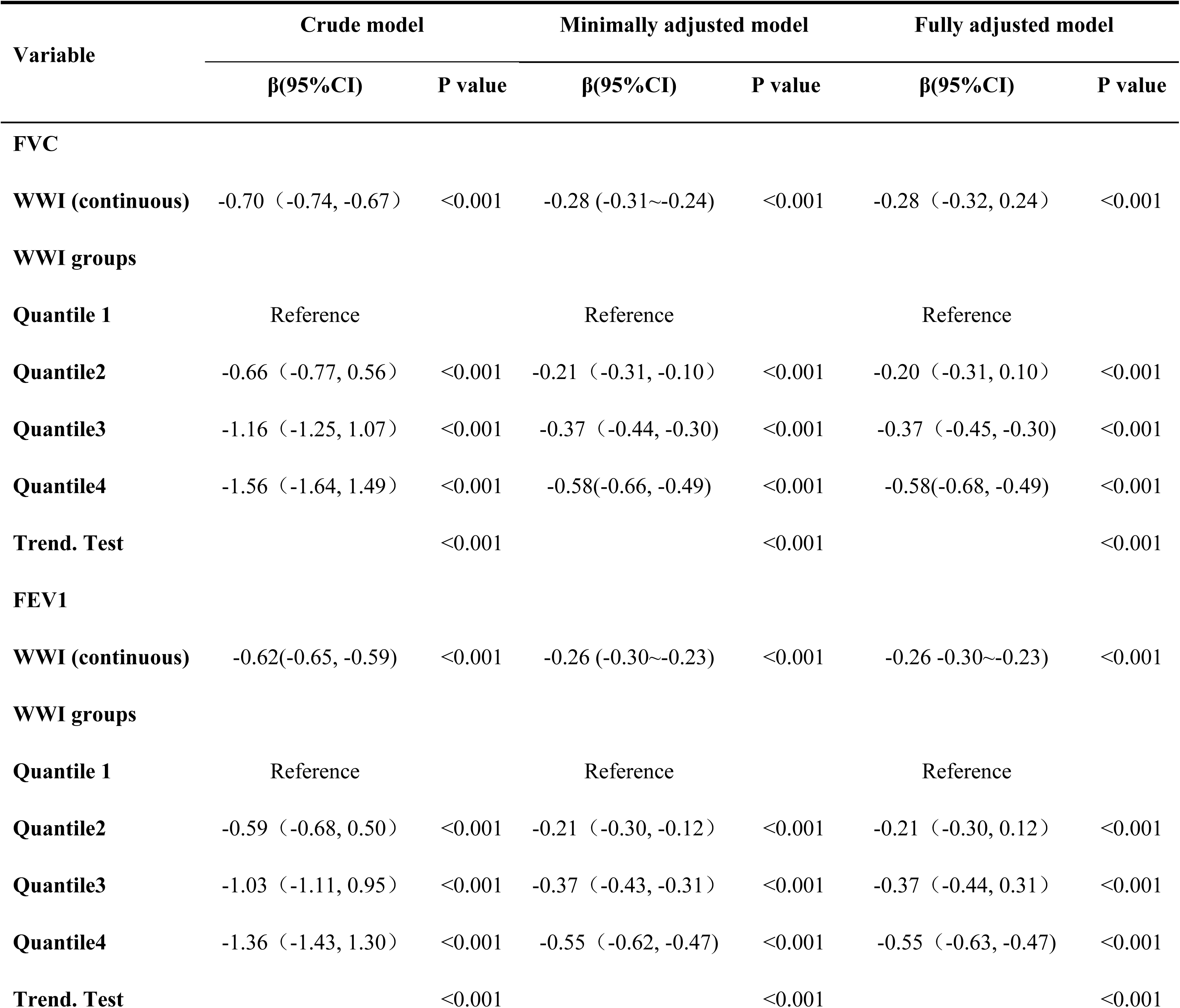
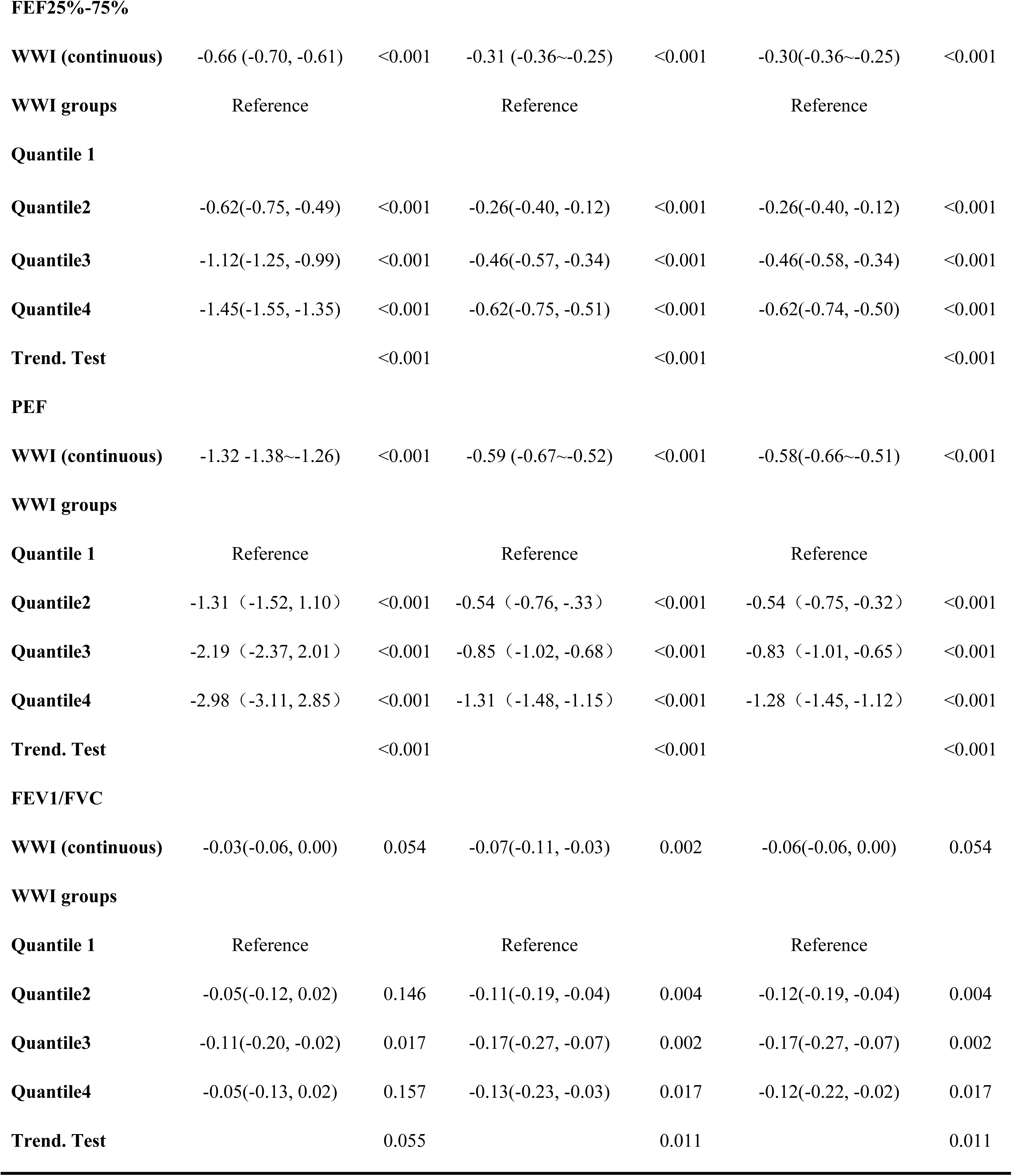
Association analysis between WWI and lung function.

| Variable | Crude model |  | Minimally adjusted model |  | Fully adjusted model |  |
| --- | --- | --- | --- | --- | --- | --- |
|  | β(95%CI) | P value | β(95%CI) | P value | β(95%CI) | P value |
| **FVC** |  |  |  |  |  |  |
| **WWI (continuous)** | -0.70 (-0.74, -0.67) | <0.001 | -0.28 (-0.31~-0.24) | <0.001 | -0.28 (-0.32, 0.24) | <0.001 |
| **WWI groups** |  |  |  |  |  |  |
| **Quantile 1** | Reference |  | Reference |  | Reference |  |
| **Quantile2** | -0.66 (-0.77, 0.56) | <0.001 | -0.21 (-0.31, -0.10) | <0.001 | -0.20 (-0.31, 0.10) | <0.001 |
| **Quantile3** | -1.16 (-1.25, 1.07) | <0.001 | -0.37 (-0.44, -0.30) | <0.001 | -0.37 (-0.45, -0.30) | <0.001 |
| **Quantile4** | -1.56 (-1.64, 1.49) | <0.001 | -0.58(-0.66, -0.49) | <0.001 | -0.58(-0.68, -0.49) | <0.001 |
| **Trend. Test** |  | <0.001 |  | <0.001 |  | <0.001 |
| **FEV1** |  |  |  |  |  |  |
| **WWI (continuous)** | -0.62(-0.65, -0.59) | <0.001 | -0.26 (-0.30~-0.23) | <0.001 | -0.26 -0.30~-0.23) | <0.001 |
| **WWI groups** |  |  |  |  |  |  |
| **Quantile 1** | Reference |  | Reference |  | Reference |  |
| **Quantile2** | -0.59 (-0.68, 0.50) | <0.001 | -0.21 (-0.30, -0.12) | <0.001 | -0.21 (-0.30, 0.12) | <0.001 |
| **Quantile3** | -1.03 (-1.11, 0.95) | <0.001 | -0.37 (-0.43, -0.31) | <0.001 | -0.37 (-0.44, 0.31) | <0.001 |
| **Quantile4** | -1.36 (-1.43, 1.30) | <0.001 | -0.55 (-0.62, -0.47) | <0.001 | -0.55 (-0.63, -0.47) | <0.001 |
| **Trend. Test** |  | <0.001 |  | <0.001 |  | <0.001 |
| **FEF25%-75%** |  |  |  |  |  |  |
| **WWI (continuous)** | -0.66 (-0.70, -0.61) | <0.001 | -0.31 (-0.36~-0.25) | <0.001 | -0.30(-0.36~-0.25) | <0.001 |
| **WWI groups** | Reference |  | Reference |  | Reference |  |
| **Quantile 1** |  |  |  |  |  |  |
| **Quantile2** | -0.62(-0.75, -0.49) | <0.001 | -0.26(-0.40, -0.12) | <0.001 | -0.26(-0.40, -0.12) | <0.001 |
| **Quantile3** | -1.12(-1.25, -0.99) | <0.001 | -0.46(-0.57, -0.34) | <0.001 | -0.46(-0.58, -0.34) | <0.001 |
| **Quantile4** | -1.45(-1.55, -1.35) | <0.001 | -0.62(-0.75, -0.51) | <0.001 | -0.62(-0.74, -0.50) | <0.001 |
| **Trend. Test** |  | <0.001 |  | <0.001 |  | <0.001 |
| **PEF** |  |  |  |  |  |  |
| **WWI (continuous)** | -1.32 -1.38~-1.26) | <0.001 | -0.59 (-0.67~-0.52) | <0.001 | -0.58(-0.66~-0.51) | <0.001 |
| **WWI groups** |  |  |  |  |  |  |
| **Quantile 1** | Reference |  | Reference |  | Reference |  |
| **Quantile2** | -1.31 (-1.52, 1.10) | <0.001 | -0.54 (-0.76, -.33) | <0.001 | -0.54 (-0.75, -0.32) | <0.001 |
| **Quantile3** | -2.19 (-2.37, 2.01) | <0.001 | -0.85 (-1.02, -0.68) | <0.001 | -0.83 (-1.01, -0.65) | <0.001 |
| **Quantile4** | -2.98 (-3.11, 2.85) | <0.001 | -1.31 (-1.48, -1.15) | <0.001 | -1.28 (-1.45, -1.12) | <0.001 |
| **Trend. Test** |  | <0.001 |  | <0.001 |  | <0.001 |
| **FEV1/FVC** |  |  |  |  |  |  |
| **WWI (continuous)** | -0.03(-0.06, 0.00) | 0.054 | -0.07(-0.11, -0.03) | 0.002 | -0.06(-0.06, 0.00) | 0.054 |
| **WWI groups** |  |  |  |  |  |  |
| **Quantile 1** | Reference |  | Reference |  | Reference |  |
| **Quantile2** | -0.05(-0.12, 0.02) | 0.146 | -0.11(-0.19, -0.04) | 0.004 | -0.12(-0.19, -0.04) | 0.004 |
| **Quantile3** | -0.11(-0.20, -0.02) | 0.017 | -0.17(-0.27, -0.07) | 0.002 | -0.17(-0.27, -0.07) | 0.002 |
| **Quantile4** | -0.05(-0.13, 0.02) | 0.157 | -0.13(-0.23, -0.03) | 0.017 | -0.12(-0.22, -0.02) | 0.017 |
| **Trend. Test** |  | 0.055 |  | 0.011 |  | 0.011 |

Statistical analyses involved three regression models. Crude model: No covariates were adjusted. The minimally adjusted model controlled for age, citizenship status, sex, race or ethnicity, and educational attainment. The fully adjusted model included all covariates: birth weight, serum cotinine level, age, citizenship status, sex, race or ethnicity, educational attainment, poverty–income ratio (PIR), and history of asthma in a blood relative. The findings are displayed as regression coefficients (β) together with P values and 95% CIs.CI: confidence interval.

### Analysis of Nonlinear Relationships and Threshold Effects

To further assess the dose-response nature of the WWI- pulmonary function link, we fitted smoothed exposure-response curves **(Fig2)**. These analyses indicated nonlinear associations between WWI and FVC, FEV1, FEF25%–75%, PEF, and the FEV1/FVC ratio. Threshold effects were subsequently assessed using two-piecewise linear regression models. This analysis identified significant inflection points (thresholds) for WWI at 10.83 (FVC), 10.83 (FEV1), 10.70 (FEF25%–75%), 11.00 (PEF), and 10.79 (FEV1/FVC ratio) **(Table 3)**. The fit of the piecewise models was significantly better than that of simple linear models (likelihood ratio test, P<0.001 for all outcomes), suggesting a change in the association slope at these WWI values.

**Table 3.**
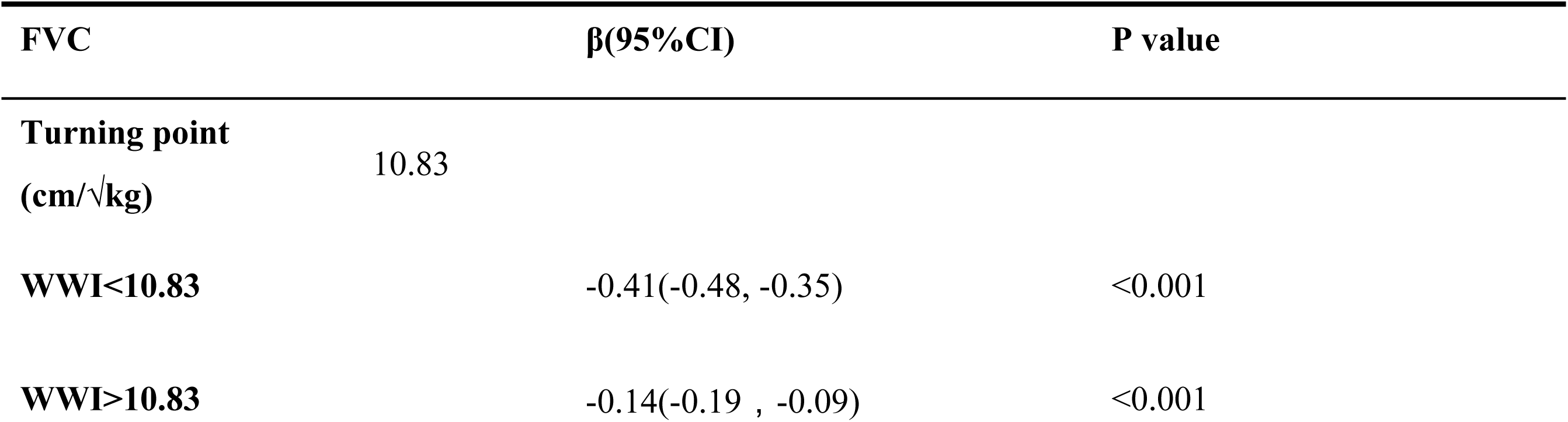
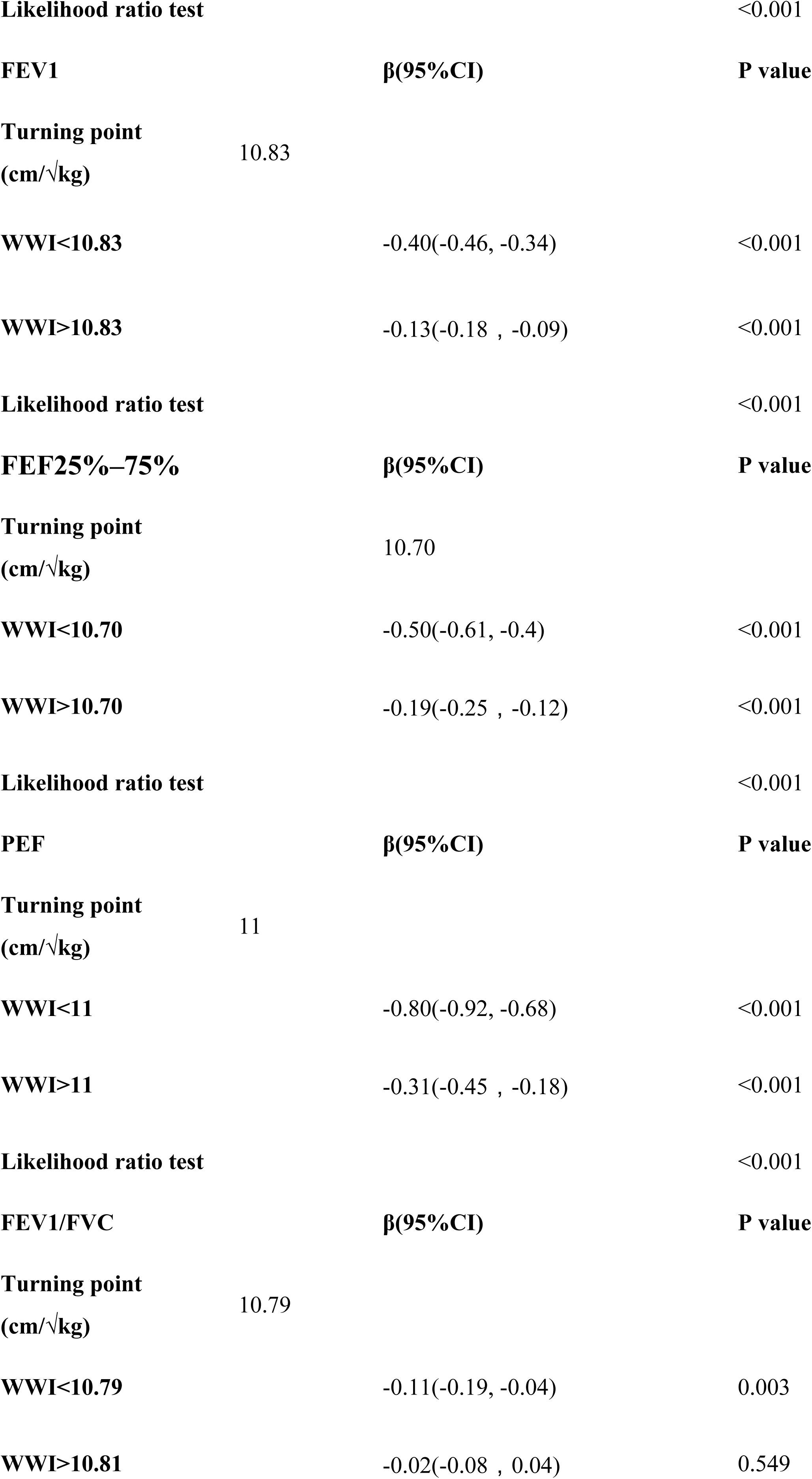
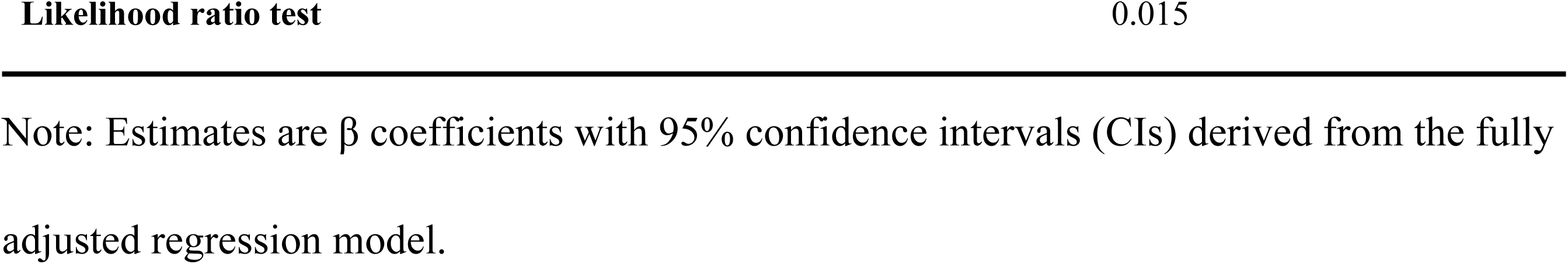
Analysis of Thresholds in the WWI–Lung Function Relationship.

### Subgroup analyses

The association between WWI and pulmonary function was further explored by stratifying the analysis according to age, sex, race or ethnic group, weight at birth, serum cotinine level and poverty-income ratio (PIR) **(Fig 3)**. A stable inverse association between WWI and Pulmonary function indicators (FVC, FEV1, FEF25%-75% and PEF) was observed across most strata, although the magnitude of association was greater among males. Specifically, stronger inverse associations were noted for: FVC: Among participants <12 years of age, males, Non-Hispanic White participants, those with serum cotinine <0.05 ng/mL (no recent tobacco exposure), and those with PIR ≥3.5.FEV1: Among males, participants identifying with other racial or ethnic groups, those with serum cotinine <0.05 ng/mL, and those with PIR ≥3.5.FEF25%–75%: Among males and participants with normal birth weight.FEV1/FVC Ratio: Among participants ≥12 years of age and those identifying with other racial or ethnic groups. These results indicate potential heterogeneity in the association between WWI and lung function across subgroups.

**Fig 3.**
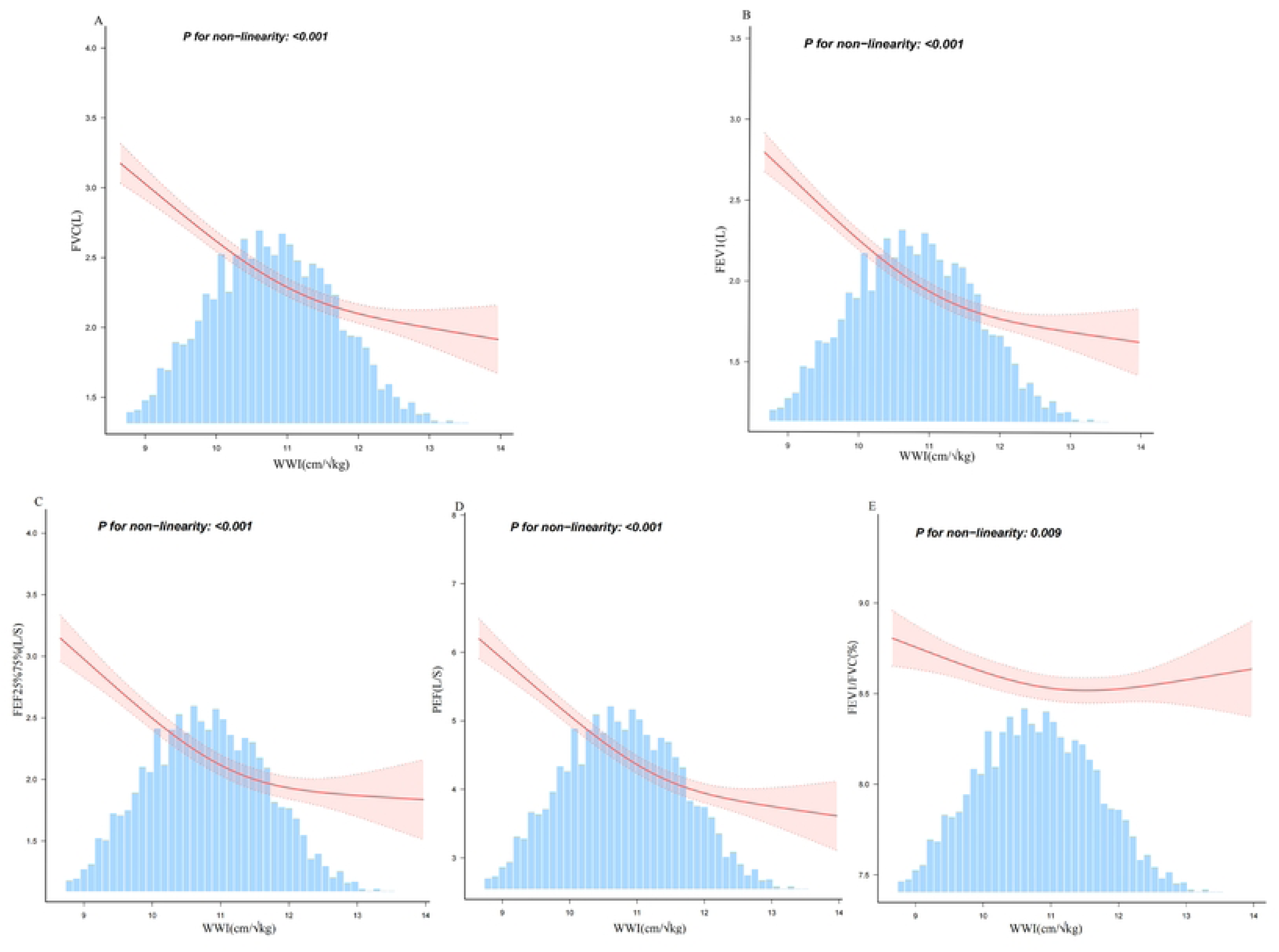
Subgroup Analysis of the Association between WWI and Pulmonary Function. Data points represent β coefficients (horizontal lines indicate 95% CI) from the fully adjusted model.

To explore potential sex differences, we performed curve fitting analyses stratified by sex.

The shape of the correlation between WWI and pulmonary function measures differed by sex **(Fig 4)**. An L-shaped relationship connected WWI with FVC, FEV1, FEF25%–75%, and PEF specifically in boys, contrasting with the findings for girls. The link between WWI and pulmonary function in children appears to vary between sexes.

**Fig 4.**
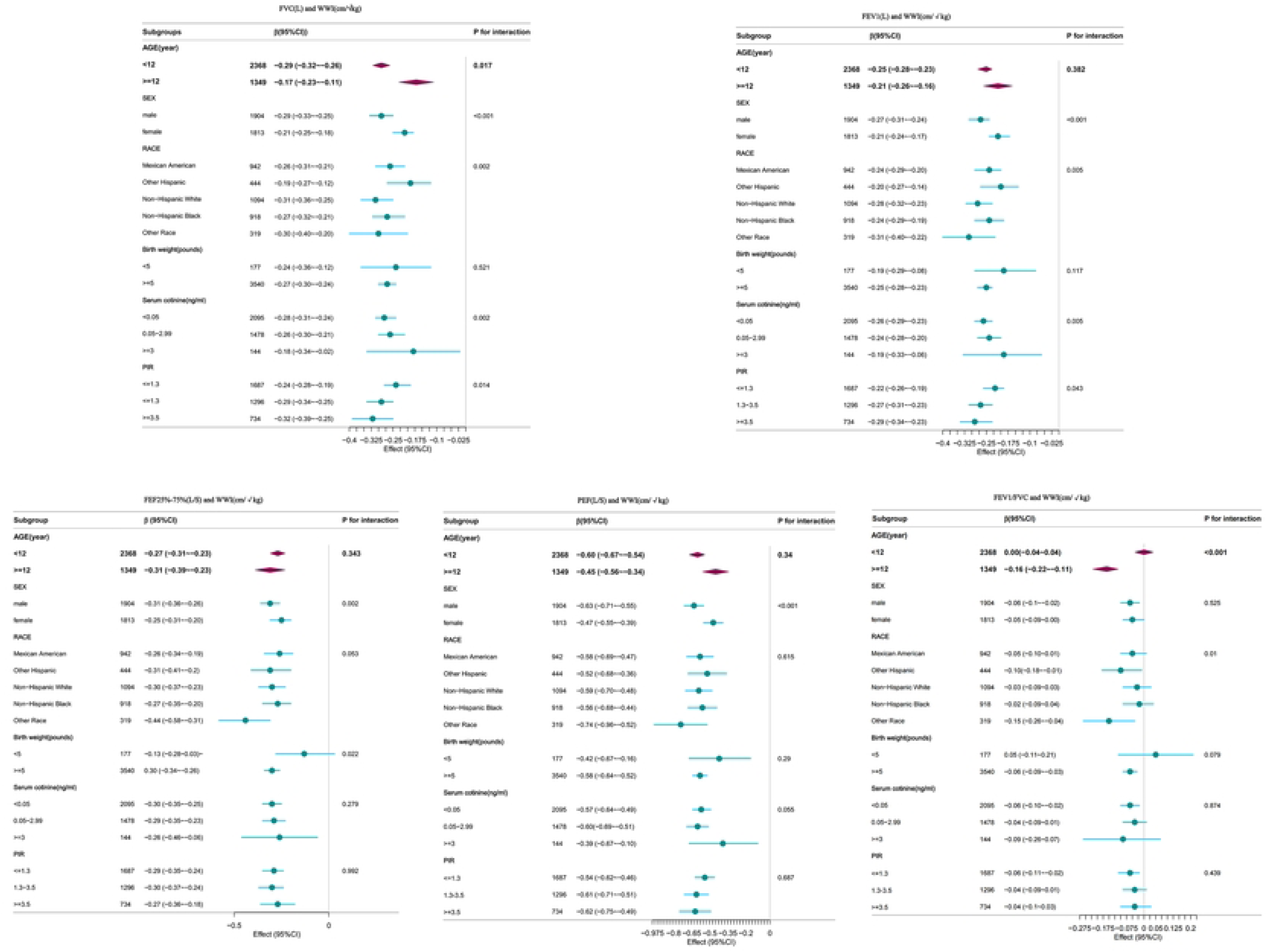
Sex-Stratified Association between WWI and Pulmonary Function. Curves represent the estimated association (shaded areas indicate 95% CI) for males and females, based on the fully adjusted model.

## Discussion

Using data from NHANES 2007–2012 for 3717 U.S. children ages (6–17) We analyzed the link between WWI and pulmonary function. We believe this relationship has not been previously assessed in a pediatric population. The results of the weighted multiple linear regression model show that higher WWI is negatively correlated with FVC, FEV1, FEF25%- 75% and PEF. Analyses using WWI quartiles (Q1–Q4) and nonlinear curve fitting consistently revealed an inverse association. Consistent inverse associations between WWI and FVC, FEV1, FEF25%–75%, and PEF were observed across strata in subgroup analyses. These links seemed stronger in boys than in girls. They indicate that the effects of WWI on pulmonary function may be more noticeable population subgroups. These findings underline the significance of focused assessment and intervention. The link between WWI and lung function appeared different for boys and girls, based on sex-stratified curve fits.

Consequently, assessing WWI’s effect on children’s lung function should take sex-specific influences into account. The use of real lung capacity measurements rather than estimated percentages and the strict control of confounding factors including age, sex, and race may be responsible for these results(28). By using this methodological technique, the estimated relationship between WWI and pulmonary function is more reliable.

Our findings suggest WWI is linked to pulmonary function in children and may be an important factor. Extensive research demonstrates a correlation between obesity and numerous chronic diseases, like diabetes, certain cancers and cardiovascular disease(29,5).Beyond these, abdominal obesity stands out as a particularly pernicious form(30), with increased visceral fat being a significant factor contributing to metabolic syndrome, which encompasses conditions like hypertension(31) and diabetes.

Musculoskeletal health is also affected by obesity, which increases the incidence of osteoarthritis, in weight-bearing joints like the hips and knees (32).Excessive mechanical stress from weight gain speeds up joint inflammation and cartilage deterioration. Moreover, obesity is a driver of Ongoing low-intensity inflammation and metabolic dysfunction, which can lead to organ complications and further health deterioration(33).

Research also highlights a significant Link between obesity and decreased lung function. In pediatric populations, research (34) reveals that higher BMI correlates with lower FEV1/FVC ratios, pointing to potential airway obstruction. A meta-analysis(6) further solidifies this connection by demonstrating reduced lung function across all metrics in obese children.

Additionally, a Chinese cohort study linked visceral fat accumulation to poorer lung function(35). Collectively, these findings point to obesity severity playing an important role in pulmonary function. Accurate assessment of obesity can facilitate early identification of children at risk for lung function impairment, offering valuable insights for preventive and therapeutic interventions.

Research on the “obesity paradox” reports that people with grade 1 obesity (BMI 30-<35) don’t have increased death rates from all causes compared to normal-weight individuals, while those who are overweight (BMI 25-<30) tend to have lower rates(36).Current research(18,37,38) indicates that traditional obesity metrics like BMI and the circumference of the waist have limitations, often failing to distinguish between muscle mass and fat. Contributing to the “obesity paradox,” this shortcoming undermines the reliability of results based on these metrics. For instance, one investigation (39) showed FVC was not significantly related to body weight or BMI. Conversely, higher waist circumference (in both sexes) and higher waist-to-hip ratio (in men) were linked to lower FVC. This suggests that measures of abdominal adiposity may associate more strongly with pulmonary function than do BMI or body weight.

Consistent with other findings, the circumference of the waist is inversely correlated with pulmonary function regardless of BMI category (normal weight, overweight, or obese), a consistency not seen with BMI(40). Moreover, research(41) on normal weight obesity (NWO) reveals an inverse association between body fat percentage and pulmonary function (FEV1 and FVC). Ruderman introduced the idea of metabolically obese normal-weight (MONW) individuals in 1981(42) .MONW(43) individuals, who have excess visceral fat despite a normal BMI (18–25 kg/m^2^), are at an increased risk of metabolic syndrome, underscoring the limitations of BMI in capturing metabolic health. These insights highlight the limitations of using traditional obesity measures to accurately predict lung function. The WWI(18), a unique index that takes abdominal fat into consideration and provides a more trustworthy option for predicting the health effects of obesity, was introduced by Park et al. WWI has been validated number of illness predictions (19,21,22)and is increasingly recognized as an obesity metric. A study (44)demonstrated that rising WWI values across ethnic groups correspond to increased abdominal fat and decreased muscle mass, with age exacerbating these trends. Thus, WWI emerges as a promising anthropometric indicator. Our study extends these observations to children, demonstrating a similar inverse association between WWI and pulmonary function measures in this population.

The specific pathways linking WWI to pulmonary function in children remain to be fully elucidated. However, several plausible hypotheses can be proposed based on current evidence. Obesity impacts lung function through multiple pathways, including mechanical stress, chronic inflammation, and metabolic alterations(45). These processes might work together to impair lung function and raise the risk of respiratory illnesses. Adipokines(46) which are secreted by adipose tissue and are engaged in intricate biological processes, constitute an active endocrine organ. The body enters a persistent, low-grade chronic inflammatory state (40,47) because of the imbalance of adipokines (increased pro-inflammatory and decreased anti-inflammatory) and changes in the internal environment of adipose tissue (hypoxia and immune cell infiltration). These two factors work together to disturb immune homeostasis. In addition to having an impact on the adipose tissue itself, this condition also impacts lung health and increases susceptibility to respiratory tract infections to directly damage lung tissue and indirectly impair lung function. Associations between biomarkers reflecting oxidative stress and decrements in pulmonary function have been observed in previous studies(48). Obesity is also closely linked to metabolic syndrome, where insulin resistance(45) can exacerbate oxidative stress, further damaging lung tissue.

Furthermore ,The distribution of body fat, particularly in the chest and abdomen(49), can mechanically restrict diaphragm movement and increase intrathoracic pressure, especially in supine positions. This may lead to alveolar collapse and premature closure of peripheral airways, affecting gas exchange. Additionally, Stratified analyses revealed varying nonlinear relationships between WWI and lung function across different sexes. These differences might be explained by how male and female immune systems respond differently; for instance, females(50) could be more susceptible to chronic low-grade inflammation when obese, potentially impacting their lung function in distinct ways.

Our study has several significant benefits. This is the first research using the WWI obesity measure to look at children’s lung function. It aims to better understand the link between obesity and lung function and explore the reasons behind it. Additionally, the use of high-quality, nationally representative data from NHANES 2007–2012 allows our findings to be applied more broadly to the U.S. population. We also meticulously accounted for sample weights and a wide array of confounders, supporting the robustness of the results. We should acknowledge certain limitations of this study. Because we used a cross-sectional design, we cannot conclude that WWI causes the observed pulmonary function differences. Investigating this relationship over time through longitudinal research is necessary to assess causality.

Additionally, while we controlled numerous covariates, residual confounding may still exist. Future research could address this by incorporating a broader set of covariates and employing advanced methods like propensity score matching or mediation analysis to strengthen causal claims.

## Conclusion

We observed an inverse association between WWI and pulmonary function among U.S. children, highlighting the potential relevance of this adiposity measure to pediatric respiratory health. Despite the unclear causal relationship, these findings warrant attention and highlight the need for further experimental and longitudinal research to solidify these conclusions.

## Supporting Information

**S1 Checklist.** STROBE Checklist for reporting cross-sectional studies.

**S1 Dataset.** Data used in the study on Weight-Adjusted Waist Index and pulmonary function in children.

## Acknowledgments

We are grateful to the National Center for Health Statistics for providing access to the NHANES database.

## Author contributions

Wenjun Zhai and Xinyuan Yang: Conceptualization, methodology, software, Writing-review & editing.

Guopeng Liang, Jiang li Chen, Yongfang Zhou: Project administration, Supervision. Wenjun Zhai: Data curation, writing original draft.

Junya Zhou: Writing – review & editing

All authors contributed to the article and approved the submitted version.

## Data availability statement

The datasets used in this study are publicly available from the National Center for Health Statistics (NCHS) through the National Health and Nutrition Examination Survey (NHANES) for the years 2007–2012. The data can be accessed at the following URL: https://www.cdc.gov/nchs/nhanes/.

## Ethics statement

All survey protocols were approved by the NCHS Ethics Review Board, and written informed consent was obtained from all participants.

